# Multivariable MR can mitigate bias in two-sample MR using covariable-adjusted summary associations

**DOI:** 10.1101/2022.07.19.22277803

**Authors:** Joe Gilbody, Maria Carolina Borges, George Davey Smith, Eleanor Sanderson

## Abstract

Genome-Wide Association studies (GWAS) are hypothesis free studies that survey the whole genome for polymorphisms associated with a trait of interest. To increase power and to estimate the direct effects of these single nucleotide polymorphisms (SNPs) on a trait GWAS are often conditioned on a covariate (such as body mass index (BMI) or smoking status). This adjustment can introduce bias in the estimated effect of the SNP on the trait. Mendelian randomisation (MR) studies use summary statistics from GWAS estimate the causal effect of a risk factor (or exposure) on an outcome. Covariate adjustment in GWAS can bias the effect estimates obtained from MR studies using the GWAS data. Multivariable MR (MVMR) is an extension of MR that includes multiple traits as exposures. Using simulations we show that MVMR can recover unbiased estimates of the direct effect of the exposure of interest by including the covariate used to adjust the GWAS within the analysis. We show that this method provides consistent effect estimates when either the exposure or outcome of interest has been adjusted for a covariate. We apply this method to estimate the effect of systolic blood pressure (SBP) on type-2 diabetes (T2D) and waist circumference on systolic blood pressure both adjusted and unadjusted for BMI.

## Introduction

Mendelian randomisation (MR) uses the special properties of germline genetic variants to strengthen inference regarding the influence of potentially modifiable exposures on disease (1, 2). MR can be implemented as an instrumental variables (IV) analysis, using the genetic variants as instruments. MR requires a set of assumptions to obtain reliable estimates of the effect of the exposure on the outcome First that the genetic variants used as instruments are associated with the risk factor of interest, secondly that there are no unmeasured confounders of the association between the genetic variants and outcome and thirdly that the genetic variants act on the outcome only through the risk factor of interest (3). A fourth assumption is required for the interpretation of the magnitude of effect sizes, assuming that there is a monotonic association between the instrument and the exposure (2, 4).

MR is often conducted using genetic variants identified through genome wide association studies (GWAS). GWAS use a hypothesis free methodology and test the association of hundreds of thousands of single nucleotide polymorphisms (SNPs) with a risk factor in large populations to identify the particular SNPs associated with that risk factor (5, 6). Most SNPs found to be associated with a trait have a relatively small effect and explain a small proportion of the heritability of the trait (7).

MR can be performed using summary data from GWAS by combining the SNP-exposure and SNP-outcome associations to estimate the causal effects of the exposure on the outcome (2, 8). As each variant explains a relatively small amount of the variation of a trait, large sample sizes are often required to have sufficient power for MR effect estimation. Current large GWAS either use large datasets like UK Biobank which contains over 500,000 individuals (9) or the very largest GWAS use groups of biobanks and have sample sizes of well over 1 million (10). However some of the largest GWAS studies available have been conditioned on a covariate aiming to improve the statistical power of the study or to estimate direct effects of the genetic variants on the trait of interest that do not act via the covariate. For example, cardiometabolic traits such as blood pressure are often adjusted for body mass index, lung function, height and smoking status, etc (11-13).

These covariate adjustments can introduce bias into the estimated SNP trait associations (14). Firstly covariate adjustment biases which SNPs are identified by the GWAS, by including SNPs associated with the covariate as well as the trait of interest. Figure 1A displays the mechanisms through which collider adjustment can cause genetic variants with no direct effects on the trait of interest to be identified as having an association with the trait of interest. The adjustment for the covariate creates a path between the genetic variant and the trait due to the unmeasured common genetic or environmental cause between the covariate and the trait. As conditioning for a collider creates a path between the genetic variant and the trait (15), it causes a non-causal association to be identified, altering the range of SNPs that would be found by the GWAS as it would no longer solely identify SNPs associated with the trait of interest. To eliminate this would require adjusting for the common cause. Secondly as shown in figure 1B, when an unmeasured genetic or environmental risk factor (G_-g_ and E) is shared between the covariate (C) and the trait of interest (X) the covariate acts as a collider in the path between the genetic variant (g) and outcome leading to biased estimates for the direct effect of the genetic variant on the trait of interest. Collider bias occurs when two variables (such as g and G_e_/E in figure 1) that independently cause a third variable (C in figure 1) are statistically adjusted for the third variable (16).

**Figure 1.**
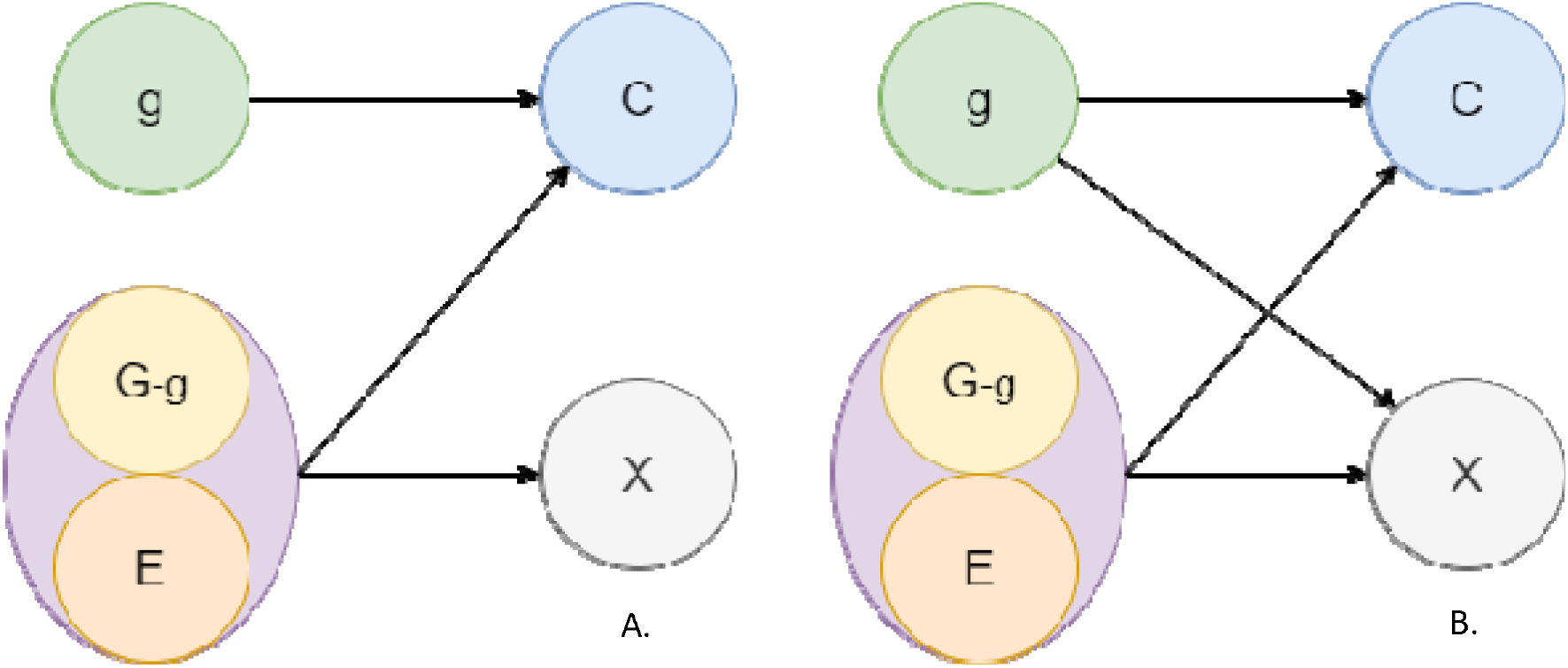
Diagrams describing the causal relationship between a genetic variant g, uncorrelated other genetic loci G_-g_, environmental factors E, covariate C and the trait of interest X. In A the correlation between C and X is explained by shared risk factors, leading to an indirect pathway between g and X while in B a direct pathway exists between g and X but is biased due to shared risk factors between C and X.

Previous simulation studies have shown that the use of covariable-adjusted GWAS in MR analyses can bias effect estimates in ways that are difficult to predict and that are highly dependent on the underlying (and unobserved) causal structure (17, 18). In simulations where no unobserved common causes of the exposure and outcome in the MR study exist the covariate adjustment eliminates bias in the presence of pleiotropic effects of the SNP on the covariate. However the existence of unobserved confounders is one of the main motivations for performing MR. In simulations where unobserved confounding was present covariate adjustment will lead to bias, particularly when there are unobserved confounders between the covariate and the outcome in the MR study (17). However, lack of alternative data means many two sample MR studies have used these adjusted summary results previously, for example while investigating the effect of adiposity or waist-to-hip ratio on cardiometabolic traits (19, 20). The use of these covariate adjusted GWAS potentially leads to biased results obtained from those studies, and at the time of writing we are not aware of a proposed solution to recover this effect.

Multivariable Mendelian randomisation (MVMR) is an extension of MR that estimates the effect of multiple exposures on an outcome (21, 22). MVMR estimates the direct effect of each exposure on the outcome conditional on the other exposures included in the estimation, rather than the total effect as obtained in univariable MR. MVMR can therefore be used to determine whether multiple exposures exert a causal effect on the outcome or one mediates the causal effect of another. MVMR can also estimate direct causal effects of multiple exposures where the genetic variants are thought to have pleiotropic effects through those exposures

Here we investigate the use of MVMR including the covariate that the GWAS for the exposure of interest was adjusted for as an additional exposure. We do this to recover an estimate of the true direct causal effect estimate of the exposure of interest on the outcome when only adjusted GWAS results are available for either the exposure or the outcome. We consider settings where either the exposure or outcome of interest have been adjusted for a covariate in the GWAS. We use simulation studies to explore the results obtained for both exposure and outcome GWAS adjustment under a range of scenarios for the relationship between the exposure and covariate. We then apply this to estimate the causal effect of systolic blood pressure (SBP) on type 2 diabetes (T2D) where the GWAS for SBP has been adjusted for BMI and to estimate the effect of waist circumference (WC) on SBP where either WC or SBP are adjusted for BMI and compare these results to those obtained for estimation using GWAS that were not adjusted for BMI.

## Simulation study

### Methods

We performed a simulation study to explore whether it is possible to recover the direct effect of an exposure of interest by including the covariate in a MVMR estimation when the GWAS for either the exposure of interest or outcome has been adjusted for the covariate.

Simulations were performed under 3 different causal structures in the data for the relationship between the exposure of interest (X_1_), the covariate (X_2_) and the outcome (Y). In each case an unobserved confounder acts on both exposures (X_1,_ X_2_) and the outcome (Y), leading to X_1_ and X_2_ being correlated even when neither has a direct effect on the other. In all simulations X_1_ and X_2_ have a causal effect on Y.

G_1_ and G_2_ are sets of genetic variants that satisfy the instrumental variable assumptions for X_1_ and X_2_ respectively. For each setting considered we conducted three simulations, once with the G_1_-X_1_ associations estimated without adjustment and once with the G_1_-X_1_ associations adjusted for X_2_ and once with the G_1_-Y associations adjusted for X_2_. In each case we also obtained estimated G_2_-X_2_ associations, without any adjustment, for use in MVMR estimation.

The first underlying structure (A1 - Confounded model) describes a situation where the exposure of interest (X_1_) and outcome (Y) are confounded by a covariate (X_2_). The second scenario (A2 - Correlated Model) describes a scenario where X_1_ and X_2_ are correlated through shared (unobserved) confounding. The final causal structure (A3 - Mediated model) describes a scenario where X_1_ is has an effect on X_2_ as well as Y, meaning X_2_ mediates the X_1_, Y relationship. These scenarios are illustrated below (Figure 2).

**Figure 2.**
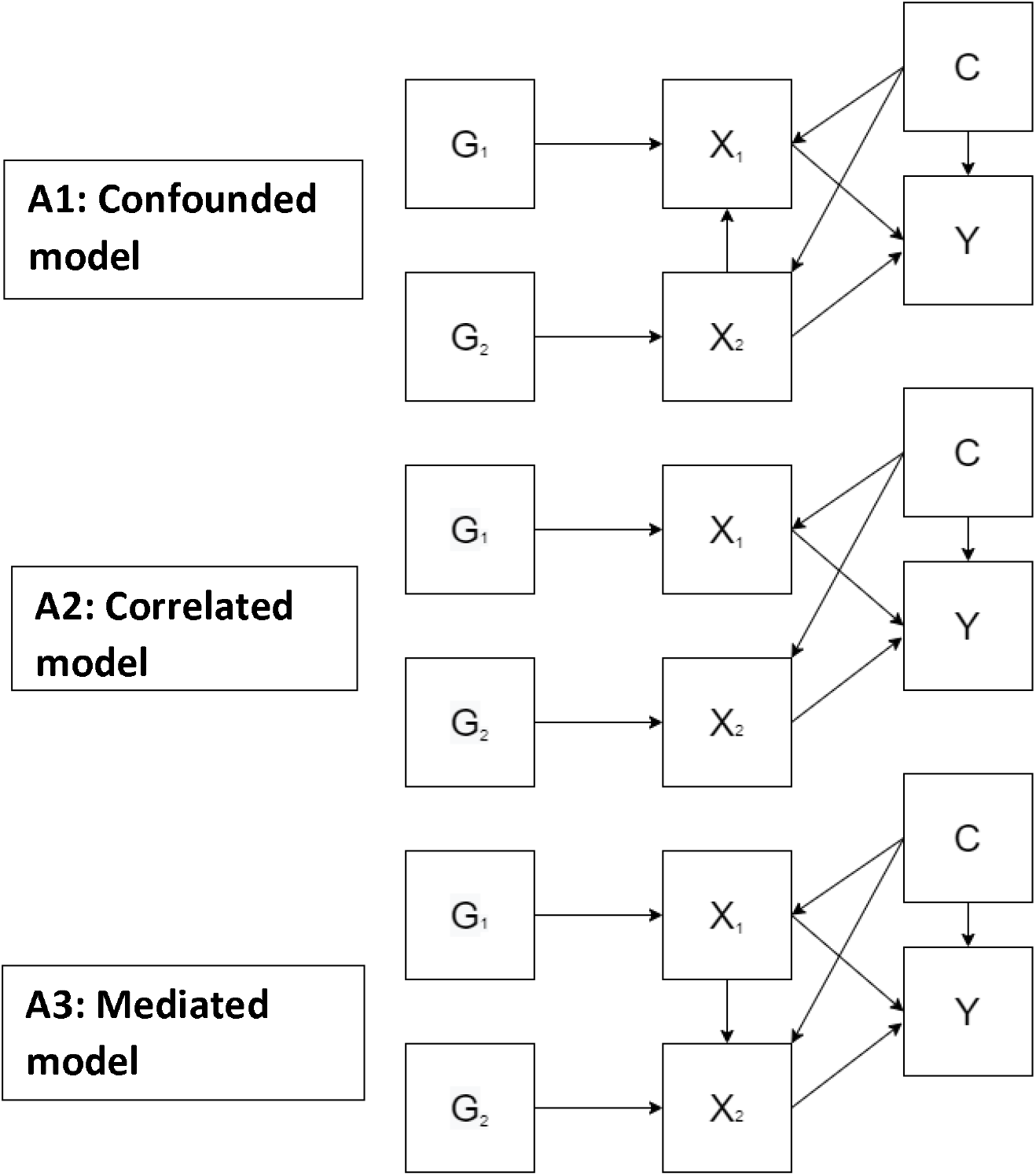
Directed acyclic graphs (DAG) charts displaying the underlying causal structures of the simulated datasets, where G represents simulated variants, X a simulated exposure, Y a simulated outcome and C an unmeasured common cause.

MR and MVMR causal estimates were estimated using inverse variance weighting (IVW). The simulated data sets were generated using 250 simulated variants, and 10,000 simulated individuals across 5000 datasets. The effects of the simulated variants on each exposure and the outcome were estimated with a linear regression with adjustment introduced by including X_2_ as a covariate in the linear regression.

### Results

Table 1 gives the IVW univariable MR estimates from our simulation. These results show estimates of the total effect of the exposure on the outcome with and without adjustment for the covariate (*X*_2_) in the GWAS for our exposure of interest (*X*_1_). Without adjustment for the covariate estimates for *β*_1_ when X_2_ is a confounder (Model A1) are biased in the direction of *β*_2_ due to the presence of direct effect from X_2_ to X_1_. This leads to apparent pleiotropy in the SNPs associated with our exposure as some of the genetic variants associated with X_2_ are detected as being associated with X_1_ and so included in the estimation. Estimates for *β*_1_ when X_2_ is correlated with X_1_ are consistent estimates of the direct and total effect of X_1_ on Y (model 2). Estimates for *β*_1_ when X_2_ is a mediator (model A3) estimate the total effect size of X_1_ on the outcome, including an indirect effect via X_2_. When the GWAS for X_1_ is adjusted for X_2_ all three models show bias and MR estimation does not estimate the direct or total causal effect of X_1_ on Y. In each case the bias is in the opposite direction to the direct effect of X_2_ on Y (*β*_2_).

**Table 1.** IVW univariable MR of simulated data where X_1_ is unadjusted and adjusted for the covariate X_2_.

|  | Unadjusted estimates |  |  |  | Adjusted Estimates |  |
| --- | --- | --- | --- | --- | --- | --- |
| | $\beta X_1$ | Standard Error | $\beta X_2$ | Standard Error | $\beta X_1$ | Standard Error |
| **Confounded model (A1)** | 0.491 | 0.053 | 0.895 | 0.011 | 0.173 | 0.074 |
| **Correlated Model (A2)** | 0.398 | 0.009 | 0.796 | 0.011 | 0.195 | 0.067 |
| **Mediated Model (A3)** | 0.597 | 0.011 | 0.834 | 0.027 | 0.214 | 0.079 |
| Simulated direct effect sizes of $X_1 = 0.4$ and $X_2 = 0.8$. Number of SNPs = 250. N = 10,000, reps = 5000. | | | | | | |

Table 2 gives MVMR estimates of the direct effect of X_1_ and X_2_ on Y with and without adjustment for X_2_ in the estimation of the SNP X_1_ association. These results give consistent estimates of the direct effect of X_1_ on Y in both the unadjusted and adjusted estimation. However when the effect of the SNPs adjusted for X_2_ are used in the MVMR the estimate obtained for X_2_ is biased.

**Table 2.** IVW MVMR estimates of simulated data where X_1_ is unadjusted and adjusted for the covariate X_2_.

|  | Unadjusted estimates |  |  |  | Adjusted Estimates |  |  |  |
| --- | --- | --- | --- | --- | --- | --- | --- | --- |
| | $\beta X_1$ | Standard Error | $\beta X_2$ | Standard Error | $\beta X_1$ | Standard Error | $\beta X_2$ | Standard Error |
| **Confounded model (A1)** | 0.394 | 0.012 | 0.792 | 0.012 | 0.395 | 0.012 | 1.01 | 0.013 |
| **Correlated Model (A2)** | 0.395 | 0.011 | 0.791 | 0.011 | 0.395 | 0.011 | 0.913 | 0.011 |
| **Mediated Model (A3)** | 0.396 | 0.012 | 0.790 | 0.012 | 0.396 | 0.013 | 0.963 | 0.012 |
| Simulated direct effect sizes of $X_1 = 0.4$ and $X_2 = 0.8$. Number of SNPs = 250. N = 10,000, reps = 5000. | | | | | | | | |

To test the effects of covariate adjustment when the true effect of either the exposure or covariate is null, *β*_1_and *β*_2_ were alternatively set to 0. When *β*_1_ = 0 the IVW MVMR accurately estimated the effect of X_2_, even when used as an adjustment for X_1_. When *β*_2_ = 0 this pattern was not repeated, the estimated effect of X_2_ was biased as in our main analyses. These results are reported in Supplementary tables 1 - 4. We additionally explored the results obtained when X_1_ and X_2_ act in opposite directions on the outcome. These results showed the same pattern of result as obtained in the main analysis and are given in Supplementary tables 5-6.

Tables 3 and 4 show the results from MR and MVMR estimation of the same models when the GWAS for the outcome, Y, is adjusted for a covariate. These results follow the same pattern to the exposure covariate adjustment results, with a bias observed in MR estimates including the adjusted outcome in the opposite direction to the directional effect size of the covariate (X_2_) on the outcome. In the MVMR an unbiased estimate of the effect size of X_1_ is obtained for both the unadjusted and adjusted outcome estimation. While a biased estimate is generated for the effect of X_2_ on the outcome when the outcome is adjusted for X_2_.

**Table 3.** IVW univariable MR estimates of simulated data where outcome Y is first unadjusted and then adjusted for the covariate X_2_.

|  | Unadjusted estimates |  |  |  | Adjusted Estimates |  |
| --- | --- | --- | --- | --- | --- | --- |
| | $\beta X_1$ | Standard Error | $\beta X_2$ | Standard Error | $\beta X_2$ | Standard Error |
| **Confounded model (A1)** | 0.503 | 0.055 | 0.891 | 0.012 | 0.136 | 0.079 |
| **Correlated Model (A2)** | 0.398 | 0.010 | 0.789 | 0.011 | 0.170 | 0.068 |
| **Mediated Model (A3)** | 0.598 | 0.011 | 0.825 | 0.025 | 0.178 | 0.081 |
| Simulated effect sizes of $X_1 = 0.4$ and $X_2 = 0.8$. Number of SNPs = 250. N = 10,000, reps = 5000. | | | | | | |

**Table 4.** IVW MVMR estimates of simulated data where outcome Y is first unadjusted and then adjusted for the covariate X_2_.

|  | Unadjusted estimates |  |  |  | Adjusted Estimates |  |  |  |
| --- | --- | --- | --- | --- | --- | --- | --- | --- |
| | $\beta X_1$ | Standard Error | $\beta X_2$ | Standard Error | $\beta X_1$ | Standard Error | $\beta X_2$ | Standard Error |
| **Confounded model (A1)** | 0.394 | 0.013 | 0.791 | 0.013 | 0.396 | 0.013 | 1.01 | 0.013 |
| **Correlated Model (A2)** | 0.395 | 0.012 | 0.787 | 0.012 | 0.395 | 0.012 | 0.910 | 0.011 |
| **Mediated Model (A3)** | 0.396 | 0.012 | 0.787 | 0.013 | 0.396 | 0.014 | 0.960 | 0.013 |
| Simulated effect sizes of $X_1 = 0.4$ and $X_2 = 0.8$. Number of SNPs = 250. N = 10,000, reps = 5000. | | | | | | | | |

**Table 5.** IVW univariable MR effect size estimates SBP and BMI on type 2 diabetes, effect size represent odds ratios per SD increase in SBP.

|  | Unadjusted estimates |  |  |  | Adjusted Estimates |  |
| --- | --- | --- | --- | --- | --- | --- |
|  | SBP |  | BMI |  | SBP (adjusted for BMI) |  |
|  | OR | 95% CI | OR | 95% CI | OR | 95% CI |
| **IVW** | 1.47 | [1.25 - 1.72] | 1.41 | [1.25 – 1.58] | 1.36 | [1.14-1.64] |
| **MR-Egger** | 1.07 | [0.66 - 1.75] | 1.36 | [0.99 – 1.86] | 1.04 | [0.65-1.65] |
| **Weighted Median** | 1.25 | [1.01 - 1.55] | 1.46 | [1.23 – 1.74] | 1.07 | [0.89-1.30] |
| **Weighted Mode** | 0.99 | [0.64 - 1.53] | 1.47 | [1.16- 2.11] | 1.09 | [0.76-1.56] |
| OR: Estimated odds ratio for Type 2 diabetes. 95% CI, 95% confidence interval for the estimated effect. Effects estimated using 244 SNPs identified for SBP and 461 for BMI, summary data used was obtained from: UK Biobank (SBP unadjusted), International Consortium of Blood Pressure (SBP adjusted) (11), 70KforT2D (type-2 diabetes) (24), UK Biobank (BMI). Standard errors displayed in brackets. |  |  |  |  |  |  |

**Table 6.** IVW MVMR effect size estimates for SBP and BMI on type 2 diabetes, effect size represent odds ratios per SD increase in SBP.

|  | Unadjusted estimates |  |  |  | Adjusted Estimates |  |  |  |
| --- | --- | --- | --- | --- | --- | --- | --- | --- |
|  | SBP |  | BMI |  | SBP (adjusted for BMI) |  | BMI |  |
|  | OR | 95% CI | OR | 95% CI | OR | 95% CI | OR | 95% CI |
| **IVW** | 1.32 | 1.10– 1.60 | 1.35 | 1.19– 1.53 | 1.34 | 1.10 – 1.62 | 1.32 | 1.12 – 1.57 |
| OR: Estimated odds ratio for Type 2 diabetes. 95% CI, 95% confidence interval for the estimated effect. Effects estimated using 424 SNPs identified across both SBP and BMI for unadjusted estimates, and 490 for adjusted estimates, summary data used was obtained from: UK Biobank (SBP unadjusted), International Consortium of Blood Pressure (SBP adjusted) (11), 70KforT2D (type-2 diabetes) (24), UK Biobank (BMI). Standard errors displayed in brackets. |  |  |  |  |  |  |  |  |

**Table 7.** IVW univariable MR estimates for effect size of Waist circumference on SBP, effect size represent an increase in SBP per SD increase in WC.

|  | Unadjusted estimates |  |  |  | Adjusted Exposure Estimates |  | Adjusted outcome estimates |  |
| --- | --- | --- | --- | --- | --- | --- | --- | --- |
|  | WC |  | BMI |  | WC (Adjusted for BMI) |  | WC (Where SBP is adjusted for BMI) |  |
| | $\beta$ | 95% CI | $\beta$ | 95% CI | $\beta$ | 95% CI | $\beta$ | 95% CI |
| **IVW** | 0.036 | 0.031 - 0.103 | 0.122 | 0.095 - 0.148 | -0.062 | -0.138 - 0.001 | -0.081 | -0.145 - -0.016 |
| **MR-Egger** | 0.126 | -0.112 - 0.363 | 0.096 | 0.0266 - 0.166 | -0.069 | -0.354 - 0.231 | -0.118 | -0.346 - 0.111 |
| **Weighted Median** | 0.077 | 0.027 - 0.128 | 0.140 | 0.113 - 0.168 | -0.036 | -0.078 - 0.005 | -0.007 | -0.046 - 0.033 |
| **Weighted Mode** | 0.103 | -0.019 - 0.225 | 0.160 | 0.105 - 0.215 | -0.042 | -0.118 - 0.034 | -0.007 | -0.063 - 0.049 |
| $\beta$: Estimated effect size for Systolic blood pressure. 95% CI, 95% confidence interval for the estimated effect. Effect sizes estimated using 42 SNPs identified for WC in unadjusted estimates, 65 in adjusted estimates and 461 for BMI. GWAS summary data was obtained from GIANT (WC), UK Biobank (BMI, SBP unadjusted), and International Consortium of Blood Pressure (SBP adjusted). | | | | | | | | |

**Table 8.** IVW MVMR estimates for the effect size of Waist circumference on SBP estimated using MVMR, effect size represent an increase in SBP per SD increase in WC.

|  | Unadjusted MVMR estimates |  |  |  | Adjusted Exposure MVMR Estimates |  |  |  | Adjusted outcome MVMR estimates |  |  |  |
| --- | --- | --- | --- | --- | --- | --- | --- | --- | --- | --- | --- | --- |
|  | WC |  | BMI |  | WC (Adjusted for BMI) |  | BMI |  | WC (SBP is adjusted for BMI) |  | BMI (SBP is adjusted for BMI) |  |
| | $\beta$ | 95% CI | $\beta$ | 95% CI | $\beta$ | 95% CI | $\beta$ | 95% CI | $\beta$ | 95% CI | $\beta$ | 95% CI |
| **IVW** | -0.016 | -0.108 – 0.075 | 0.139 | 0.059 – 0.218 | -0.028 | -0.09 – 0.034 | 0.125 | 0.096 – 0.154 | -0.012 | -0.094 – 0.071 | -0.013 | -0.855 – 0.059 |
| $\beta$: Estimated effect size for Systolic blood pressure. 95% CI, 95% confidence interval for the estimated effect. Effect sizes estimated using 473 SNPs identified across both WC and BMI for unadjusted estimates and 474 for adjusted estimates. GWAS summary data was obtained from GIANT (WC), UK Biobank (BMI, SBP unadjusted), and International Consortium of Blood Pressure (SBP adjusted). | | | | | | | | | | | | |

These results show how adjustment for a covariate will bias the results obtained from univariable MR when SNP-trait associations for either the exposure or the outcome are obtained from covariate adjusted GWAS. MVMR including the adjustment covariate corrected for this bias and so recovered a consistent estimate of the direct effect of the exposure of interest (X_1_). The results obtained for the estimated direct effect of the covariate (X_2_) on the outcome in these models were biased and cannot be interpreted.

## Application

### Methods

To illustrate this method we considered two applications using traits that are often adjusted for body mass index (BMI). We estimate, the causal effect of systolic blood pressure (SBP) on type-2 diabetes (T2D) and of waist circumference (WC) on SBP, using both MR and MVMR with GWAS summary results. For our first example we considered adjustment of the exposure (SBP) for BMI, in the second example we consider adjustment of either the exposure (WC) or the outcome (SBP) for BMI.

To estimate the effect of SBP on T2D we used summary data for BMI and SBP from an unadjusted GWAS extracted from UK Biobank and hosted on OpenGWAS (23) and from a GWAS study adjusted for BMI using data from UK Biobank and the International Consortium of Blood Pressure-Genome Wide Association Studies (ICBP) (11). GWAS results for T2D, unadjusted for BMI, were obtained from the 70KforT2D GWAS due to lack of overlap with UK Biobank (24).

To estimate the effect of WC on SBP we used GWAS summary data for WC from a GIANT study, this data was available both unadjusted and adjusted for BMI (12). We used the same unadjusted SBP GWAS summary-statistics as in the first example (23) a BMI GWAS summary statistic were obtained from the GIANT cohort (25).

The causal effects were estimated using inverse variance weighting (IVW), genetic variants were selected on the basis that they were robustly associated with the exposure in a GWAS of that exposure, based on a genome wide significant p-value < 5e-08. Each study applied its own quality control, details of which can be found in the original publications (11, 12, 23, 25, 26). Genome-wide significant SNPs were clumped to remove SNPs in LD with each other (R2 threshold for considering LD was set to 0.001). Summary association data for the exposure and outcome for the selected SNPs were harmonised to reflect the same effect alleles and palindromic SNPs with intermediate allele frequencies were excluded using the R package ‘TwoSampleMR’ (27) and MVMR estimation with the R package ‘MVMR’ (28). For MR estimation we also conducted MR-Egger (29), Weighted Mode (30) and Weighted median (31) estimation in order to assess how sensitive the results were to potential pleiotropy. F statistics were calculated using the R^2^ statistic according to the formula as 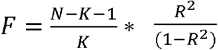 where N is the sample size of the study and K is the number of SNPs (6).

### Results

In the IVW univariate MR analyses of SBP on T2D using unadjusted summary statistics gives a positive causal effect of SBP on T2D (OR increase in T2D of 1.47 per SD increase in SBP, 95% CI: 1.25 - 1.72). The mean F statistic for the SNPs used in this analysis on SBP was 17.9. Using SBP GWAS results adjusted for BMI this effect attenuated (OR: 1.36 95% CI: 1.14-1.64). While the mean F statistic for the SNPs used in this analysis on SBP was 13.1. Our unadjusted MVMR results however show a lower direct effect than estimated for the unadjusted MR results (log OR: 1.32 95% CI: 1.10– 1.60). However the estimated effect did not attenuate when the adjusted GWAS for SBP was used (OR 1.34 95% CI: 1.10 – 1.62). Our univariable MR estimates estimated increased BMI increases risk of T2D (OR increase in T2D of 1.41 per SD increase in BMI, 95% CI: 1.25 – 1.58). This estimate size attenuated in the unadjusted MVMR estimation. Unlike the simulation results above the effect of BMI in the MVMR using the adjusted SBP has similar effect size but increased standard error to that of the unadjusted estimate.

Sensitivity analyses for the MR estimation showed inconsistent results for both unadjusted and adjusted SBP. This potentially reflects pleiotropic effects due to BMI confounding the SBP-T2D association and SNPs associated with BMI primarily being detected in the unadjusted SBP GWAS and incorrectly adjusted for in the adjusted GWAS results.

These same MVMR analyses were repeated instead using only using the SNPs identified as being associated with SBP in the SBP GWAS (11) in order to test whether the differences between the obtained results were due to differing numbers of SNPs (Supplementary tables 7-8). The estimates using both the unadjusted and adjusted sample data were very similar to the full SNP analyses. This follows the pattern set by the full SNP set analyses, in that the MVMR estimates the SBP-T2D causal effect more consistently across adjusted and unadjusted data.

For our second example we replicated the analyses above to estimate the effect of WC on SBP. The effect of waist circumference on SBP was estimated as a small increase in SBP (0.0359 mmHg per SD increase in WC, 95% CI: 0.031 - 0.103) using summary data unadjusted for the covariate BMI. The mean F statistic for these SNPs was 70.7. When using GWAS results for WC adjusted for BMI a slight decrease in SBP due to WC was estimated (−0.062 95% CI: -0.138 - 0.001). The mean F statistic for these SNPs was 45.7. Using GWAS results for the outcome SBP adjusted for BMI the estimated effect was negative like the estimates produced by the analyses using a covariate adjusted exposure (- 0.081 95% CI: -0.145 - -0.016). Here the MR Egger, weighted median and weighted mode estimates had roughly the same effect size and magnitude as the IVW estimates across all analyses. The MVMR estimates for the effect of waist circumference was a consistent decrease in SBP across all 3 analyses.

The effect size of BMI on SBP was also estimated firstly with IVW in a two sample MR setting, then in a MVMR setting including unadjusted waist circumference GWAS summary data and finally including summary data already preadjusted for BMI. All three estimates gave a similar positive estimates and sensitivity analyses for the univariable MR estimates also gave largely consistent results.

To reflect the simulated studies where the outcome is adjusted for the covariate an outcome GWAS adjusted for BMI was used, these estimates mirrored the direction of effect of the exposure adjusted results and the effect size (−0.081 mmHg per SD increase in WC, 95% CI: : -0.150 - -0.016) and (−0.012 95% CI: -0.094 – 0.071) respectively for two-sample and MVMR estimates.

These analyses were repeated using only the SNPs identified for the trait of interest (waist circumference (20)) in order to identify whether the mismatch in number of SNPs in two sample MR and MVMR analyses (45 vs 473) was unduly influencing the results of the WC effect size estimate (Supplementary tables 9-10). The unadjusted MVMR estimates for the WC only using SNPs identified for WC did not provide evidence of effect (0.176 mmHg per SD increase in WC, 95% CI: -0.999 – 1.35) the adjusted similarly estimates a null effect (−0.152 95% CI: -0.429 – 0.124). All of these analyses confidence intervals overlap with the estimates generated with the full set of SNPs.

## Discussion

Covariate adjusted GWAS results can bias the results from MR studies when either the exposure or outcome have been adjusted for a covariate (17, 18). The results from our simulation and applied analyses highlight and illustrate these biases. Here we show that MVMR can be used to obtain unbiased direct causal effect estimates from covariate adjusted GWAS study by including the covariate as an additional exposure. The estimated results obtained are the direct effect of the exposure on the outcome, conditional on the covariate. However, when used in this way MVMR cannot be used to provide an accurate estimate of the causal effect of the covariate on the outcome.

Using simulated data we found there were expected differences in IVW estimates across unadjusted two sample MR estimates of different simulated causal structures presented. When the covariate was a confounder of the exposure and outcome relationship we also observed bias because of pleiotropic effects of genetic variants associated with that covariate being included in the unadjusted MR estimation. When the covariate was a mediator the total effect estimated in the unadjusted univariable estimates was higher than the direct effect due to the effect of the mediator on the outcome. When using the adjusted summary statistics the two-sample MR IVW estimated effect of the exposure of interest was biased in the opposite direction to the true effect of the covariate on the outcome (Tables 1 and supplementary table 5). This was as expected due to the bias in the SNPs exposure association introduced by the adjustment, creating spurious associations between SNPs and the exposure and an inverse relationship between adjusted exposure and SNPs associated with the covariate. When analysing the same simulated data sets in MVMR the estimates obtained were consistent even with the covariate adjustment (Tables 1,2,3,4, and supplementary tables 5 and 6).

This same pattern of observation was repeated across the real data applied examples from our SBP-T2D example, where the two sample estimates show a difference between the causal effect estimate of the adjusted and unadjusted in the opposite direction to that of the effect direction of the covariate (BMI) on the outcome. The variation in the effect estimates obtained by the sensitivity analyses suggests that substantial horizontal pleiotropy was present. The MVMR estimate for both adjusted and unadjusted summary data were very similar, well within the 95% confidence intervals. As with our findings displayed here previous observational and MR studies have identified a positive correlation and associations between systolic blood pressure and type 2 diabetes (32, 33).

Our second applied example (WC – SBP) displayed the same pattern of results. With the two-sample estimates of the models adjusted and unadjusted for BMI being in opposite directions while the MVMR estimates were very similar in both magnitude and direction. Previous research on the relationship between these two traits finds a positive relationship between the two independent of BMI (12, 34) matching our findings.

Our study proposes a simple solution to the issue of bias in MR estimation from covariate adjustment in the GWAS. MVMR with summary level data is already widely implemented across a range of applications and therefore existing data sources and software can be easily extended to use this method. However, an important limitation of our proposed solution is that this approach can only estimate the direct effect of the exposure of interest on the outcome. This effect may differ from the total effect if the covariate is a mediator of the exposure outcome relationship and our proposed method does not provide an approach to recover the total effect of the exposure on the outcome. Estimation also requires summary GWAS results for the covariate to be available.

Another limitation of this method is that the estimate of the direct effect of the covariate on the outcome obtained from this approach is potentially biased and cannot therefore be interpreted in the same way as a conventional MVMR analysis would. The only exception to this is when the exposure has no true effect on the outcome. Further work is required to determine if the level of bias of this estimate can be calculated and so a reliable causal effect estimate calculated.

Finally, this method requires that all of the standard assumptions of MVMR, such as no weak instruments, also hold. These assumptions can be tested in the same way as any standard MVMR estimation (28, 33). However, if the exposure and covariate are closely related, or one of the adjusted exposure and covariate have only a few SNPs associated with them then it is more likely that the MVMR will be biased by weak instruments. Consequently this method of correction cannot be applied.

In the case where a choice between using a non-adjusted GWAS and an adjusted GWAS with the methodology laid out in this paper is available the non-adjusted GWAS is still preferable. Firstly, due to the ability to estimate total effect size with two-sample MR without bias due to the adjustment being introduced. Secondly when estimating effect sizes using MVMR with a non-adjusted GWAS the effect size estimates from the other exposures can be interpreted, something not possible when including the covariate in MVMR with an adjusted GWAS.

In conclusion, when using covariate adjusted summary association results from a GWAS the bias that the covariate introduces may be overcome by using a MVMR approach where the covariate used to adjusted the GWAS is included in the analysis as a second exposure.

## Data Availability

All data produced are available online at https://gwas.mrcieu.ac.uk/ and code is available at https://github.com/eleanorsanderson/adjustedGWAS

## Ethical approval

All data analysed were from publicly available summary statistics generated using relevant ethical approval from their respective studies.

## Code availability

Code for the simulation study and analysis is available at https://github.com/eleanorsanderson/adjustedGWAS v1.0

## Author Contributions

JG conducted the simulation study and applied analysis and wrote the first draft of the manuscript. ES developed the methodology and supervised the work. All authors reviewed and edited the manuscript.

