## Supplementary tables 1-10 for "Multivariable MR can mitigate bias in two-sample MR using covariable-adjusted summary associations"

**Supplementary data.**

**Supplementary Table 1. IVW univariable MR estimates of simulated data where X_1_ estimate is unadjusted and adjusted for the covariate X_2_**

|  | **Unadjusted estimates** | | | | **Adjusted Estimates** | |
| --- | --- | --- | --- | --- | --- | --- |
|  | **β X_1_** | **Standard Error** | **β X_2_** | **Standard Error** | **β X_2_** | **Standard Error** |
| **Confounded model** | 0.055 | 0.033 | 0.502 | 0.0065 | -0.119 | 0.037 |
| **Correlated Model** | -0.011 | 0.0065 | 0.502 | 0.00722 | -0.1139 | 0.0372 |
| **Mediated Model** | 0.138 | 0.00637 | 0.4864 | 0.00848 | -0.034 | 0.03719 |
| Simulated direct effect size of X1 is 0 and X2 is 0.5. Number of SNPs = 250. N = 10,000, reps -= 5000. | | | | | | |

**Supplementary Table 2. IVW MVMR estimates of simulated data where X_1_ estimate is unadjusted and adjusted for the covariate X_2_**

|  | **Unadjusted estimates** | | | | **Adjusted Estimates** | | | |
| --- | --- | --- | --- | --- | --- | --- | --- | --- |
|  | **β X_1_** | **Standard Error** | **β X_2_** | **Standard Error** | **β X_1_** | **Standard Error** | **β X_2_** | **Standard Error** |
| **Confounded model** | -0.00046 | 0.0076 | 0.499 | 0.00756 | -0.00021 | 0.0075 | 0.4992 | 0.0076 |
| **Correlated Model** | -0.013 | 0.0072 | 0.500 | 0.0074 | -0.01311 | 0.00734 | 0.4963 | 0.0078 |
| **Mediated Model** | 0.0184 | 0.0076 | 0.4828 | 0.0083 | 0.01922 | 0.00767 | 0.4912 | 0.0081 |
| Simulated direct effect size of X1 is 0 and X2 is 0.5. Number of SNPs = 250. N = 10,000, reps -= 5000. | | | | | | | | |

**Supplementary Table 3. IVW univariable MR estimates of simulated data where X_1_ estimate is unadjusted and adjusted for the covariate X_2_**

|  | **Unadjusted estimates** | | | | **Adjusted Estimates** | |
| --- | --- | --- | --- | --- | --- | --- |
|  | **β X_1_** | **Standard Error** | **β X_2_** | **Standard Error** | **β X_2_** | **Standard Error** |
| **Confounded model** | 0.499 | 0.008 | 0.12385 | 0.0065 | 0.4293 | 0.0229 |
| **Correlated Model** | 0.500 | 0.00698 | -0.00366 | 0.00649 | 0.4593 | 0.0142 |
| **Mediated Model** | 0.487 | 0.00784 | 0.0463 | 0.0330 | 0.4470 | 0.019 |
| Simulated direct effect size of X1 is 0.5 and X2 is 0. Number of SNPs = 250. N = 10,000, reps -= 5000. | | | | | | |

**Supplementary Table 4. IVW MVMR estimates of simulated data where X_1_ estimate is unadjusted and adjusted for the covariate X_2_**

|  | **Unadjusted estimates** | | | | **Adjusted Estimates** | | | |
| --- | --- | --- | --- | --- | --- | --- | --- | --- |
|  | **β X_1_** | **Standard Error** | **β X_2_** | **Standard Error** | **β X_1_** | **Standard Error** | **β X_2_** | **Standard Error** |
| **Confounded model** | 0.4996 | 0.0076 | -0.0056 | 0.00735 | 0.4999 | 0.00787 | 0.2680 | 0.0076 |
| **Correlated Model** | 0.497 | 0.0072 | -0.001271 | 0.00714 | 0.4978 | 0.00734 | 0.1546 | 0.0075 |
| **Mediated Model** | 0.488 | 0.0076 | -0.00816 | 0.00728 | 0.488 | 0.00762 | 0.2057 | 0.00721 |
| Simulated direct effect size of X1 is 0 and X2 is 0.5. Number of SNPs = 250. N = 10,000, reps -= 5000. | | | | | | | | |

**Supplementary Table 5. IVW univariable MR estimates of simulated data where X_1_ is unadjusted and adjusted for the covariate X_2_**

|  | **Unadjusted estimates** | | | | **Adjusted Estimates** | |
| --- | --- | --- | --- | --- | --- | --- |
|  | **β X_1_** | **Standard Error** | **β X_2_** | **Standard Error** | **β X_2_** | **Standard Error** |
| **Confounded model (A1)** | 0.3024 | 0.05302 | -0.6964 | 0.007470 | 0.5239 | 0.04172 |
| **Correlated Model (A2)** | 0.3984 | 0.006687 | -0.7959 | 0.008138 | 0.5459 | 0.04894 |
| **Mediated Model (A3)** | 0.1992 | 0.006108 | -0.7512 | 0.0272 | 0.5158 | 0.05129 |
| Simulated direct effect sizes of X_1_ = 0.4 and X_2_ = -0.8. Number of SNPs = 250. N = 10,000, reps -= 5000. | | | | | | |

**Supplementary Table 6. IVW MVMR estimates of simulated data where X_1_ estimate is unadjusted and adjusted for the covariate X_2_**

|  | **Unadjusted estimates** | | | | **Adjusted Estimates** | | | |
| --- | --- | --- | --- | --- | --- | --- | --- | --- |
|  | **β X_1_** | **Standard Error** | **β X_2_** | **Standard Error** | **β X_1_** | **Standard Error** | **β X_2_** | **Standard Error** |
| **Confounded model (A1)** | 0.3992 | 0.007401 | -0.7947 | 0.007703 | 0.3987 | 0.007377 | -0.5713 | 0.0125 |
| **Correlated Model (A2)** | 0.3993 | 0.007997 | -0.793 | 0.007785 | 0.3990 | 0.007988 | -0.6704 | 0.008336 |
| **Mediated Model (A3)** | 0.3991 | 0.008049 | -0.793 | 0.0123 | 0.3982 | 0.008036 | -0.6200 | 0.007712 |
| Simulated direct effect sizes of X_1_ = 0.4 and X_2_ = -0.8. Number of SNPs = 250. N = 10,000, reps -= 5000. | | | | | | | | |

**Supplementary Table 7. IVW two-sample MR estimates of SBP and BMI on Type 2 diabetes using only SNPs found to be associated with SBP, effect size represent odds ratios per SD increase in SBP**

|  | **Unadjusted estimates** | | | | **Adjusted Exposure Estimates** | | | |
| --- | --- | --- | --- | --- | --- | --- | --- | --- |
|  | **SBP** | | **BMI** | | **SBP (adjusted for BMI)** | | **SBP** | |
|  | **OR** | **95% CI** | **OR** | **OR** | **95% CI** | **OR** | **OR** | **95% CI** |
| **IVW** | 0.0359 | 0.0307 - 0.103 | 0.0823 | -0.0542 - 0.220 | -0.0615 | -0.138 - 0.00115 | -0.111 | --0.492 - 0.271 |
| β: Estimated effect size for Systolic blood pressure. 95% CI, 95% confidence interval for the estimated effect. Effect sizes estimated using 122 SNPs identified for SBP, , summary data used was obtained from: UK Biobank (SBP unadjusted), International Consortium of Blood Pressure (SBP adjusted) (8), 70KforT2D (type-2 diabetes) (16), UK Biobank (BMI). | | | | | | | | |

**Supplementary Table 8. IVW MVMR estimates of SBP and BMI on type 2 diabetes using only SNPs found to be associated with SBP, effect size represent odds ratios per SD increase in SBP**

|  | **Unadjusted estimates** | | | | **Adjusted Estimates** | | | |
| --- | --- | --- | --- | --- | --- | --- | --- | --- |
|  | **SBP** | | **BMI** | | **SBP (adjusted for BMI)** | | **BMI** | |
|  | **OR** | **95% CI** | **OR** | **95% CI** | **OR** | **95% CI** | **OR** | **95% CI** |
| **IVW** | 1.33 | 1.09-1.60 | 0.74 | 0.39 – 1.40 | 1.26 | 1.01 – 1.57 | 0.55 | 0.30 – 1.01 |
| OR: Estimated odds ratio for Type 2 diabetes. 95% CI, 95% confidence interval for the estimated effect. Effects estimated 284 SNPs identified for SBP, summary data used was obtained from: UK Biobank (SBP unadjusted), International Consortium of Blood Pressure (SBP adjusted) (8), 70KforT2D (type-2 diabetes) (16), UK Biobank (BMI). . | | | | | | | | |

**Supplementary Table 9. IVW two-sample MR estimates of waist circumference and BMI on SBP using only SNPs found to be associated with waist circumference, effect size represent increase in SBP per SD increase in WC**

|  | **Unadjusted estimates** | | | | **Adjusted Exposure Estimates** | | | |
| --- | --- | --- | --- | --- | --- | --- | --- | --- |
|  | **WC** | | **BMI** | | **WC (Adjusted for BMI)** | | **BMI** | |
|  | **β** | **95% CI** | **β** | **95% CI** | **β** | **95% CI** | **β** | **95% CI** |
| **IVW** | 0.0359 | 0.0307 - 0.103 | 0.0823 | -0.0542 - 0.220 | -0.0615 | -0.138 - 0.00115 | -0.111 | --0.492 - 0.271 |
| β: Estimated effect size for Systolic blood pressure. 95% CI, 95% confidence interval for the estimated effect. Effect sizes estimated using 42 SNPs identified for unadjusted WC and 65 for adjusted, GWAS summary data was obtained from GIANT (WC), UK Biobank (BMI, SBP unadjusted), and International Consortium of Blood Pressure (SBP adjusted). | | | | | | | | |

**Supplementary Table 10. IVW MVMR estimates of waist circumference and BMI on SBP using only SNPs found to be associated with waist circumference , effect size represent increase in SBP per SD increase in WC**

|  | **Unadjusted estimates** | | | | **Adjusted Estimates** | | | |
| --- | --- | --- | --- | --- | --- | --- | --- | --- |
|  | **WC** | | **BMI** | | **WC (adjusted for BMI)** | | **BMI** | |
|  | **β** | **95% CI** | **β** | **95% CI** | **β** | **95% CI** | **β** | **95% CI** |
| **IVW** | 0.176 | -0.999 – 1.35) | -0.115 | -1.375 – 1.150 | -0.152 | -0.429 – 0.124 | -0.076 | -1.270 – 1.12 |
| β: Estimated effect size for Systolic blood pressure. 95% CI, 95% confidence interval for the estimated effect. Effect sizes estimated using 10 SNPs identified for unadjusted WC and 12 for adjusted GWAS summary data was obtained from GIANT (WC, BMI), UK Biobank ( SBP unadjusted), and International Consortium of Blood Pressure (SBP adjusted). | | | | | | | | |
